# Cognitive Outcomes of the At-Home Brain Balance Program

**DOI:** 10.1101/2024.06.24.24309434

**Authors:** Rebecca Jackson, Yue Meng

**Author notes:** Corresponding author: Rebecca Jackson Chief Program Officer Brain Balance, 1320 Illinois Route 59, Suite 110, Naperville, IL 60563.

## Abstract

Accessibility to developmental interventions for children and adolescents could be increased through virtual, at-home delivery of home-based training programs incorporating technology. Virtual childhood training programs and their effects on cognitive outcomes in children and adolescents with developmental delays have not been well studied. To that end, this study examined the effects of the virtual at-home Brain Balance^®^ (BB) program on the cognitive task performance of children and adolescents with baseline developmental and attentional difficulties (aged 4-17 years). The at-home BB program is delivered through: (1) a computer-based format utilizing multimodal program activities previously studied in-center (multisensory stimulation with gross motor, coordination, balance, and timing activities, along with nutritional recommendations); and (2) the BB app (visual motor, auditory and visual processing, and rhythm and timing training) — creating a comprehensive program experience delivered remotely. Cognitive performance was measured by six online cognitive assessments from Creyos Health before and after 3 months of participation in the at-home BB program (N = 316) or in-center BB program (N = 4,232), compared to controls. Results showed that overall cognitive assessment scores (including attention, response inhibition, and working memory) improved after participation in either the at-home or the in-center program, compared to controls. Importantly, significant improvements over the controls were observed for two tasks, Double Trouble and Feature Match, in both programs. Further, two analyses support that the effects on cognitive performance from either delivery format, in-center or at home, are comparable in magnitude. This research: (1) presents new findings demonstrating improved cognitive performance after 3 months of the at-home BB program; (2) replicates previous findings of cognitive improvements after 3 months of the in-center BB program; and (3) suggests that the cognitive effects of virtual at-home BB training are similar to those observed for in-center BB training. Overall, the results demonstrate the effectiveness of the at-home BB program in improving cognitive functioning in pediatric populations with preexisting developmental and attentional difficulties. With virtual delivery and ease of use, at-home programs have the potential to increase access to much-needed developmental and cognitive support, ultimately reaching populations that may otherwise lack easy access to high-quality, evidence-based developmental programs.

## Introduction

Healthy cognitive development allows children and adolescents to carry out complex higher-order mental processes associated with executive functions, attention, learning, memory, and reasoning. In turn, these childhood cognitive skills significantly influence performance in academic areas such as math, reading, and writing (Bull et al, 2008; Coldren, 2013; Hajovsky et al, 2018; Re et al, 2016; Rennie et al, 2014; Taub et al, 2008). Cognitive functioning has been shown to be negatively affected in children that struggle with attention, including those with attention-deficit/hyperactivity disorder (ADHD) as well as in children with subthreshold symptoms of ADHD that do not meet the full diagnostic criteria (Hong et al, 2014; Dörrenbächer & Kray, 2019; Mogg & Bradley, 2016). For example, children with ADHD demonstrate deficits in sustained attention, response inhibition, processing speed, and working memory, compared with typically developing children (Dörrenbächer & Kray, 2019; Moura et al, 2019; Norman et al, 2017; Suskauer et al, 2008). Cognition has also been shown to be negatively impacted in mental health concerns including anxiety and depression (Pickerring et al, 2022; Umadevi et al, 2019; Wang et al, 2023; McCarty et al, 2007).

Although pharmacological treatment is often the first-line option for managing the symptoms of inattention in many conditions including ADHD and autism, concerns surrounding long-term use, side effects, and compliance in children warrant the need to explore nonpharmacologic approaches (Perwien et al, 2004; Storebø et al, 2018). Numerous studies suggest that cognitive abilities during development can be improved through various types of training and practice (Bediou et al, 2018; Christiansen et al, 2019; Klingberg et al, 2005; Oei & Patterson, 2013; Posner et al, 2015; Tang et al, 2007), pointing to the value of multimodal approaches to support cognitive development during childhood and adolescence (Alderman et al, 2017).

One such multimodal training program (the Brain Balance^®^ program) has been shown to significantly improve overall cognitive performance as well as performance on distinct tests of attention and concentration, memory, reasoning, and verbal ability in children and adolescents who participated in the center-based Brain Balance program for three months (Jackson & Wild, 2021). In another study, center-based Brain Balance participants experienced on average a decrease in ADHD symptoms in parent-rated scores on the Brown Attention-Deficit Disorder Scales^®^ (Jackson & Jordan, 2022). More than half of these youth participants experienced statistically significant reliable change in attention from pre- to post-program, particularly in those with more pronounced attentional issues at baseline. Similarly, an open exploratory study reported that children with ADHD who completed the Brain Balance program for 15 weeks experienced improvement in total ADHD scores including reductions in the subscores for inattentive and hyperactivity on both parent- and clinician-rated measures, compared to typically developing controls (Teicher et al., 2023).

In addition to the above mentioned effects of Brain Balance participation on cognition and attention, center-based participation has demonstrated improvements on aspects of mental well-being (Jackson & Robertson, 2020), and developmental outcomes (Jackson & Jordan, 2023). Specifically regarding developmental outcomes, improvements were reported in primitive reflex integration and sensory motor development (measured in the pre and post-program assessment), as well as statistically significant reliable change in reading/writing, academic engagement, emotionality, behavior, and social communication, as reported by parents, in Brain Balance participants from pre- to post-program, with the probability and degree of change increasing as the participants’ baseline severity increased (Jackson & Jordan, 2023). All of the published studies on the center-based Brain Balance program have been conducted in children and adolescents (ages 4-17 years) with baseline attentional and developmental challenges, suggesting the potential of the Brain Balance program as a nonpharmacological approach to addressing attentional and cognitive difficulties in youth with preexisting developmental issues.

Preliminary findings suggest that the effects of the center-based Brain Balance program could be replicated in a school-based setting. In a study conducted on-site at a school where professionally trained Brain Balance staff implemented the program protocols, there were demonstrated improvements in classroom attention and behavior as reported by teachers, notably a reduction in teacher-rated scores of inattention and hyperactivity/impulsivity (Jackson & Glanz, 2023). Like the center-based studies, this school-based study was also conducted with children and adolescents with baseline attention and developmental challenges.

The Brain Balance program operates as an in-center program at various locations across the United States. However, access to these center locations may be limited by the ability of some families to travel to a center-based program or by a lack of centers where the families reside. The public health emergency resulting from the COVID-19 pandemic also presented unprecedented opportunities to develop innovative virtual programs for children using digital therapeutic and communication technologies. In order to address the need for virtual or remote programs for children and adolescents, Brain Balance created and implemented a virtual at-home program, which was designed based on the established center-based Brain Balance program involving regular frequency and duration of training. The at-home program utilizes a digital therapeutic (the proprietary Brain Balance app) that includes standardized activities related to visual motor skills, auditory and visual processing, and rhythm and timing, in addition to physical exercises and the use of specialized sensory gear, allowing for the virtual delivery of a consistent program across varying locations.

Within the healthcare and medicine space, a virtual service-delivery model is not new. A large and growing body of literature supports leveraging digital therapeutics and telecommunication technologies to expand the reach of healthcare providers to pediatric patients who would otherwise face barriers to healthcare access (Burke et al., 2015; Fung & Ricci, 2020; Gloff et al., 2015; K. M. Myers et al., 2007; Rabatin et al., 2020; Sharma et al., 2020). It is predicted that telehealth will likely become part of our ongoing model of care (Burke et al., 2015; Rabatin et al., 2020). Further, the feasibility and efficacy of providing digital therapeutics and telehealth have been demonstrated in pediatric populations including those with significant medical conditions and complex healthcare needs (Beani et al., 2020; Cady et al., 2008; Chen et al., 2013), as well as those with psychiatric disorders (K. Myers et al., 2015; K. M. Myers et al., 2007; Sharma et al., 2020). However, a virtual app-based delivery of non-medical training programs for children and adolescents with developmental and attentional issues has not been well studied.

There is emerging evidence that digital therapeutics including mobile apps could be a promising means for strengthening cognitive skills (Bonnechère et al., 2021; Leung et al., 2021; Pappas & Drigas, 2019). This study aimed to evaluate cognitive performance in children and adolescents before and after training in the at-home virtual Brain Balance program, which incorporates the app as a digital therapeutic as an element in a comprehensive program to assess whether the effects are similar to the improvements already observed within the in-center program. To this end, we retrospectively reviewed cognitive outcomes of children and adolescents with baseline developmental and attentional difficulties following three months of participation in either the at-home or center-based Brain Balance program, compared to controls. Users of the Brain Balance app (combined with additional sensory and physical activities) are trained and supported through a telehealth style program coach and a program portal with videos, instructions, and support resources. The combination of professionally trained Brain Balance coaches, access to program gear/equipment, a program portal with videos and support resources, and the Brain Balance app creates a comprehensive remote program experience that can scale beyond the limitations of a physical center-based location.

The results presented here confirmed our previous findings on cognitive outcomes in center-based Brain Balance participants (Jackson & Wild, 2021), in a larger sample size than what was used in the previous study. We found that in-center participants demonstrated improvements in aspects of cognition including attention, response inhibition, short-term and working memory, attention and concentration, and visual-spatial reasoning and strategy. Further, the results demonstrated improvements in cognitive outcomes following virtual at-home Brain Balance participation, as measured by web-based cognitive testing batteries. Cognitive gains from the virtual at-home Brain Balance program were measured in the same key areas as for the in-center program, including attention, response inhibition, short-term and working memory, and visual-spatial reasoning and strategy. We also showed that cognitive scores from virtual participation were not significantly different from in-center participation, with participants demonstrating improvement regardless of the location of program delivery. These findings suggest that consistent participation in the Brain Balance program — whether in-center or a virtual at-home setting — may serve as a potential nonpharmacologic alternative or adjunct to supporting cognitive development, including attention, response inhibition, and memory in children with developmental difficulties and attentional issues.

## Methods

### Ethical Approval

Approval for this retrospective data review was granted by an institutional review board (IRB) at Advarra (Columbia, Maryland, USA), an independent organization accredited by the U.S. Office for Human Research Protections and the Association for the Accreditation of Human Research Protection Programs. The Advarra IRB determined that this retrospective data review met the requirements for exemption from IRB oversight, according to the Department of Health and Human Services regulations found at 45 CFR 46.104(d)(4). Informed parental consent was obtained for any participants prior to general enrollment in the Brain Balance program.

### Data Source

We retrospectively reviewed archived cognitive assessment data measured by Creyos Health (formerly known as Cambridge Brain Sciences) from students enrolled at Brain Balance center locations across the United States and who met the inclusion criteria described below. Data were derived from pre- and post-program assessments collected between 2019 and 2023 on a total of 16,330 participants (1,354 at-home Brain Balance and 14,976 in-center Brain Balance participants). All participants were between the ages of 4 and 17 years (Mean [M] = 9.64, Standard Deviation [SD] = 3.18) and 67.8% were male.

### Measures

Cognitive testing was administered before and after 3 months of participation in the Brain Balance program to look for changes in cognitive functioning associated with virtual at-home and in-center Brain Balance training. Testing took place via the web-based testing platform from Creyos Health, which has previously been used for numerous large-scale studies of cognitive performance (Hampshire et al., 2012; Nichols et al., 2020; Owen et al., 2010; Stafford et al., 2020; Wild et al., 2018).

Participants completed six cognitive tests, for which detailed descriptions (including screenshots and test-retest reliability) can be found in the supplementary materials of Wild et al (2018).

Briefly, the following were the tasks used: 1) Spatial Span (short-term memory); 2) Double Trouble (a modified Stroop task measuring attention and response inhibition); 3) Monkey Ladder (visuospatial working memory); 4) Rotations (ability to manipulate and understand a visual representation); 5) Feature Match (measure of attention and concentration); and 6) Token Search (working memory and strategy). Each cognitive task has a primary outcome measure that best reflects overall performance for that task (Wild et al, 2018), and our results are based on this measure unless otherwise specified.

### Inclusion Criteria

Prior to enrolling in the Brain Balance program, prospective students were evaluated by trained assessors from Brain Balance who had completed training in the centers’ protocols. Students who were eligible for enrollment in the Brain Balance program did not have any known genetic disorders and tested below age-appropriate levels in functional measurements including visual motor, auditory and visual processing, balance, coordination, and rhythm and timing. Students also needed to demonstrate a developmental readiness for the program, as defined by the ability to engage with instructors and follow a one-step direction, to attempt the tasks requested, and to continue to work throughout the duration of the assessment. Re-direction and repetition of instructions both visually and verbally were allowed in the definition of readiness. At the time of this pre-program assessment, students completed all six Creyos cognitive tasks. Students who met the above mentioned inclusion criteria were then enrolled for participation in the Brain Balance program, as described in more detail in the *Training Protocol* section below. Following completion of the Brain Balance program, participants again completed the same Creyos cognitive tasks.

### Training Protocols

The training protocols for the virtual and in-center programs differed in the following ways:

1. The frequency of sessions – while the in-center program involved three sessions per week at the center (plus daily home exercises, as described further below), the virtual at-home program consisted of five sessions per week without additional activities to supplement the sessions. The rationale for assigning more sessions per week for the virtual program was to help ensure the same number of repetitions of exercises as the in-center program.
2. The in-center program activities were overseen by a Brain Balance coach for every session. For the virtual at-home program, participants received a virtual coach-led session once per week to evaluate the participants’ levels for each task and make appropriate adjustments to ensure that each participant is working at their challenge point in all activities. During this session, the coach also provided answers and support for parents’ questions on implementing the program for the remaining four parent-led sessions per week. Parents were provided with live instruction on the exercises, as well as videos for each program level, written descriptions, and tracking tools to help implement the program from home.
3. Certain activities from the in-center program were omitted from the virtual at-home program due to differences in equipment available, including a measure of post-rotary nystagmus, the Purdue fine motor task, balance exercises on a balance beam, and the Interactive Metronome task for rhythm and timing (virtual participants instead used the Rhythmicity task in the Brain Balance app).
4. For the in-center program, parents were asked to assist their children in completing daily exercises at home, which consisted of 0-8 primitive reflexes (assigned if the primitive reflex was present at the time of assessment), physical fitness activities (push-ups and sit-ups), and eye strengthening exercises. To ensure consistency in parental implementation of the parent-supported portions of the program, parents received training on how to perform the exercises and were provided access to an online parent portal that included videos on each of the exercises as well as written instructions with photos. These exercises are incorporated into the five day a week session schedule for consistency of program activities and quantity of repetitions across the two modes of program delivery.
5. The program structure differed for the virtual at-home versus the in-center program, in that new exercises were introduced each week in the at-home program so that families were not required to learn all program exercises the first week. The program builds progressively each week so that all exercises were introduced within the first 6 weeks of the program. The at-home program recommends completing the activities 5 days per week, so that the number of repetitions of exercises completed are the same as in the in-center program consisting of three days per week of exercises. Both program delivery methods include exercises and activities that are progressive in nature and change in duration, quantity, and complexity as the participants’ functional abilities improve over the course of the program.

More specifically, the at-home Brain Balance program consisted of the following exercises and activities:

- Passive sensory stimulation in the form of tactile, olfactory, visual, and auditory stimulation (Woo et al., 2015);
- Exercises targeting primitive and postural reflexes (Chandradasa & Rathnayake, 2020), which were assigned based on indicators of a retained reflex at the time of the initial assessment. The following reflexes were assessed: Moro reflex, spinal galant reflex, rooting reflex, palmar grasp reflex, asymmetrical tonic neck reflex, symmetrical tonic neck reflex, tonic labyrinthine reflex, and Landau reflex;
- Core muscle exercises (Myer et al., 2011);
- Proprioceptive and balance training, using a rocker board and one-leg balance (Fong et al., 2016; Kobel et al., 2020);
- Gait exercises, including agility activities, and using the cross-crawl march (Surburg & Eason, 1999) and jump rope (Trecroci et al., 2015);
- Rhythm and timing exercises completed at-home using Rhythmicity (UCSF Neuroscape, San Francisco, CA), an app-based game designed to assess sensorimotor synchronization ability with auditory, visual, and/or tactile stimuli (Johnson et al., 2020);
- Activities that aim to enhance auditory and visual processing, as well as coordination and endurance of eye movements (Fisher et al., 2015; Robert et al., 2014). More specifically, auditory engagement consisted of exposure to varying levels of auditory stimulation and activities targeting the ability to filter and rapidly process auditory information. Visual stimulation was achieved through exposure to color and light stimulation, as well as exercises that require eye coordination, timing, and speed of processing perceived information. Visual motor activities were completed using the Brain Balance app allowing all participants to complete the same exercises, including those related to gaze stabilization, saccade touch, convergence, the optokinetic reflex, visual attention, and visual processing.

The academic component of the sessions was based on the initial functional assessment and focused on improving literacy and listening skills. Nutritional guidance was provided for all participants in the in-center and virtual at-home programs.

### Analysis

To assess the effects of the Brain Balance program on cognitive functioning, including the specific areas of cognition that demonstrated significant improvements, Brain Balance participants were administered cognitive testing through the Creyos web-based testing platform before and after the program. The assessment consisted of a collection of six cognitive tasks in each test.

#### Participants

The Creyos data set contained cognitive test data for 1,408 participants who had been enrolled in the at-home Brain Balance program. Of those participants, 316 completed 3 months of the Brain Balance program and also completed pre- and post-program cognitive assessments within that time frame. The Creyos data set also contained cognitive test data for 15,499 participants of the in-center Brain Balance program. Additionally, there were 124 children who qualified for the Brain Balance program and completed the Creyos cognitive tests at least twice but did not choose to enroll. After removing subjects with reported ages older than 18 years or younger than 3 years, there were 1,354 participants who completed the at-home Brain Balance program and 14,976 participants who completed the in-center program, as well as 118 in the group that qualified for the program but did not enroll.

Next, for both the at-home and in-center programs, participants were assigned to either a control (CTRL) group or a treatment (Brain Balance [BB]) group. Two types of participants were assigned to the CTRL group: those who were enrolled and had less than 30 days between two cognitive test dates, and those who were not enrolled but tested at least twice. Participants assigned to the BB group consisted of those who were enrolled in a BB program and completed at least 3 months of training.

For the at-home BB program, the BB group consisted of 310 participants and the CTRL group consisted of 189 participants, among which 75 were enrolled and 118 were not enrolled.

For the in-center BB program, the BB group had 4,166 participants and the CTRL group had 373 participants, among which 255 were enrolled and 118 were not enrolled. Note that the same 118 non-enrolled participants were included in the CTRL group for both program delivery types.

#### Data analysis

Data were analyzed using custom code written in Python and R using open-source packages including NumPy, Pandas, Statsmodels, dplyr, rstatix, and ggplot2. Outliers defined as more than four standard deviations away from the mean test scores were removed on a per-task basis. Finally, test scores were standardized to have a mean of 0 and standard deviation of 1.0 for each cognitive task (*z* scores).

First, two-tailed Welch’s *t*-tests were used to evaluate whether the BB group and the CTRL group in the at-home and in-center programs have the same mean age at the time of the first cognitive test. Welch’s *t*-test is robust to unequal sample sizes and variance between groups, which makes it appropriate here because the BB group versus CTRL group comparison has unequal sample sizes. Gender ratio between the BB group and the CTRL group was assessed using Chi-square tests. Second, cognitive test score data for the at-home and the in-center programs were analyzed separately with a three-way mixed design repeated measures ANOVA. Treatment group (BB, CTRL) was entered as a between-subject factor, and cognitive task (six tasks: SS, Spatial Span; DT, Double Trouble; ML, Monkey Ladder; RT, Rotations; FM, Feature Match; TS, Token Search) and time point (pre-test or post-test) were entered as within-subject factors. Levene’s test was used to confirm homogeneity of variance for the between-subject factor at each level of the within-subject factors. Sphericity was assessed with Mauchly’s test; Greenhouse-Geiser correction was applied when the sphericity assumption was violated.

Lastly, to evaluate whether the treatment effects differ between the at-home and the in-center programs, we initially used three-way repeated measures ANOVA to assess whether program type had an impact on cognitive test scores of the two BB groups. In this analysis, only standardized test scores from the at-home and the in-center BB groups were used. Similar to previous ANOVA designs, cognitive task and time point were entered as within-subject factors. Instead, the between-subject factor was program type (at-home, in-center). Because we hypothesized that the treatment effect of the two programs does not differ, to support our conclusion from the aforementioned repeated measures ANOVA, bootstrapping was used to further test whether raw change scores from the six cognitive tasks were significantly different between the at-home BB group and the in-center BB group. Raw pre-test to post-test change scores from both programs were randomly resampled with replacement 10,000 times, with sampling size equal to the original group size. This was done separately for the six cognitive tasks. Each pair of bootstrapped sample means forms an at-home:in-center ratio. The proportion of the times that the at-home mean is greater than the in-center mean, or the at-home mean is smaller than the in-center mean, whichever is smaller, is considered to be the *p*-value that the two sample means are unequal in the bootstrapped two-sample hypothesis testing.

## Results

For the at-home program, there was no difference in age (*t*(406.92) = -0.65, *p* = 0.51, BB age mean = 10.33, SD = 3.15, CTRL age mean = 9.99, SD = 3.12) or distribution of males and females (*χ*(1) = 1.84, *p* = 0.17) between the BB group and the CTRL group. The BB group and the CTRL group from the in-center program had slightly different mean ages (*t*(433.50) = 4.02, *p* < 0.001, BB age mean = 10.00, SD = 3.09, CTRL age mean = 10.44, SD = 3.24), but did not differ in gender ratio (*χ*(1) = 0. 29, *p* = 0.59).

**Figure 1.**
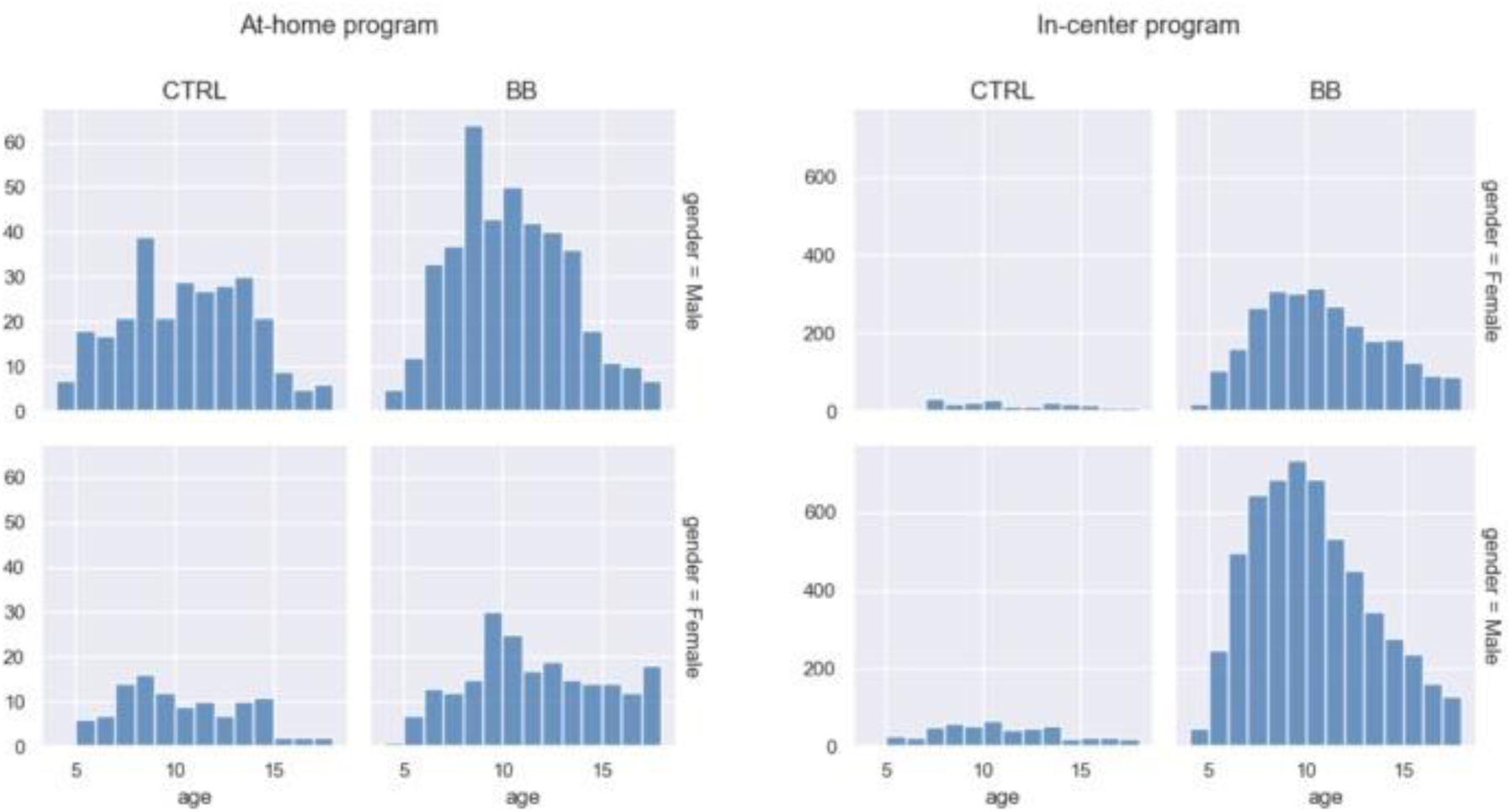
Age distribution for the two genders and treatment groups. Data from the at-home program and in-center program are plotted on separate panels.

For the at-home program, the homogeneity of variance assumption was not met under some levels, but because ANOVA is robust to such violations when group sizes are close, we proceeded with the analysis. The cognitive test data from the at-home program revealed a main effect of time point (*F*(1, 322) = 16.41, *p* < .001, ***η***2 = 0.048) and a main effect of cognitive task (*F*(1, 598.94) = 9.26, *p* < .001, ***η***2 = 0.028). There was also a significant interaction between time point and treatment group (F(1, 322) = 8.69, *p <* .01,***η***2 = 0.026).

To follow up the two-way interaction, we computed simple main effects of the treatment group by fixing time point to pre-test and post-test. At pre-test, scores between the BB and the CTRL groups did not differ (F(1, 2428) = 0.18, p = .67, ***η***2 = .000074). At post-test, the BB group outperformed the CTRL group (F(1, 2523) = 25.3, p < .001, ***η***2 = .01) (Fig. 2). Specifically, the BB group did better than the CTRL group in Double Trouble (*F*(1, 396) = 4.33, *p* < .05, ***η***2 = .011), Feature Match (*F*(1, 472) = 6.02, *p* < .05, ***η***2 = .013) and Spatial Span (*F*(1, 481) = 9.24, *p* < .01, ***η***2 = .019) (Fig. 3).

**Figure 2.**
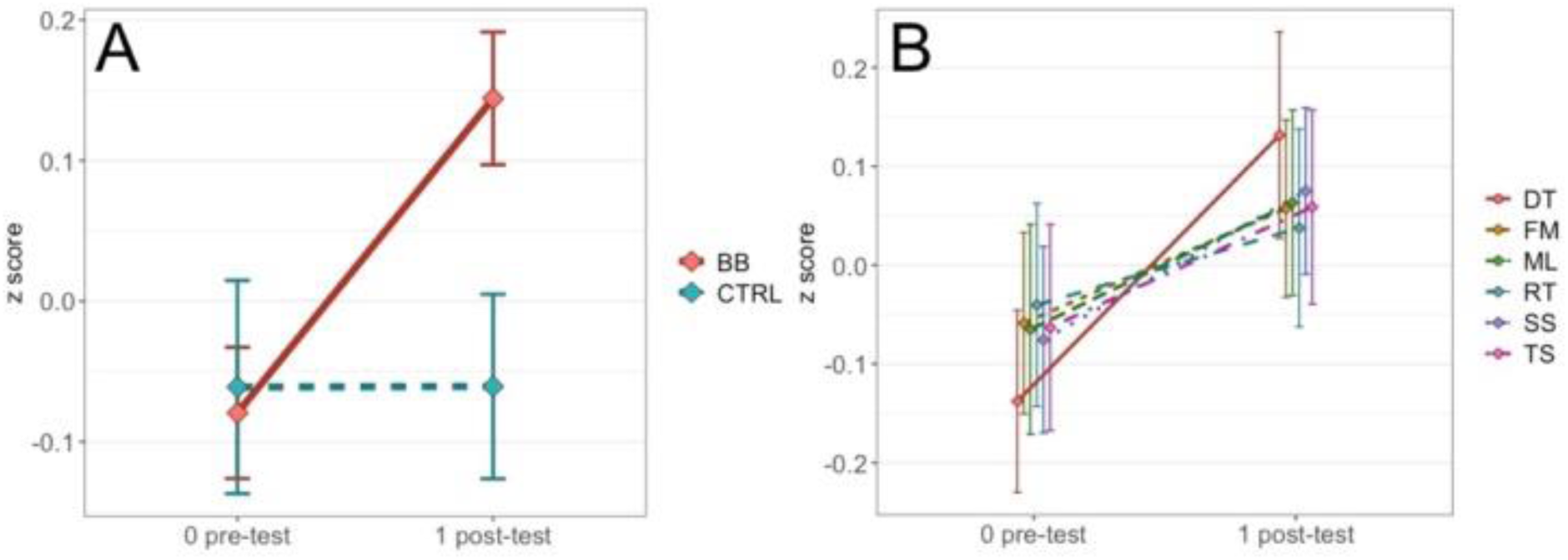
A) Comparing performance in pre- and post-tests between the BB group and the CTRL group for the at-home program. B) The time point-by-test name interaction is not significant for the at-home program, but the plot is shown as a contrast to results for the in-center program reported in the Results.

**Figure 3.**
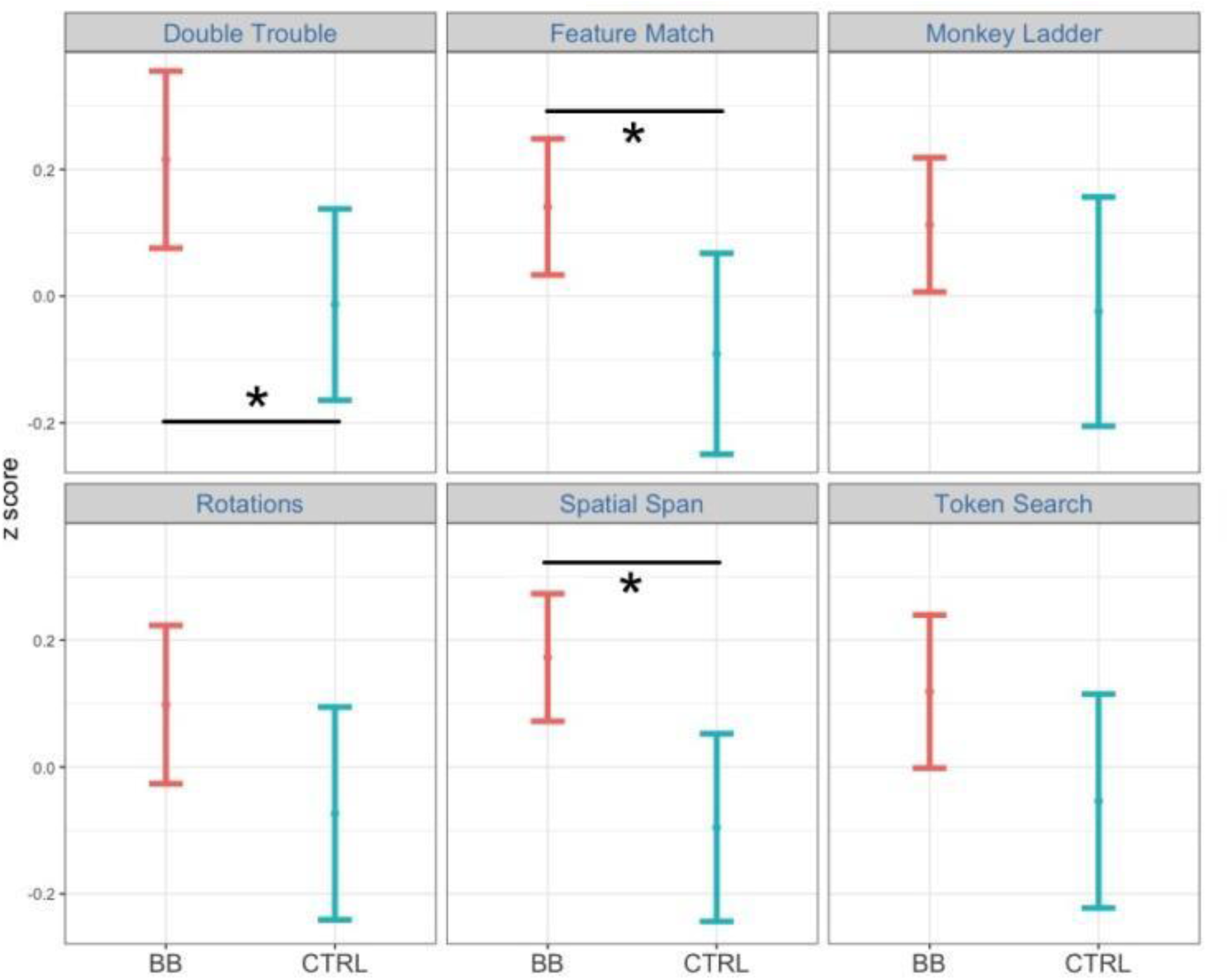
Comparing performance between the BB group and the CTRL group at post-test for the at-home program. Among the six tasks, participants in the BB group outperformed those in the CTRL group in Double Trouble, Feature Match, and Spatial Span.

For the in-center data, Levene’s test was not significant for all other conditions except for the post-test scores of the FM task. The in-center program had a significant main effect of treatment group (*F*(1, 2792) = 4.80, *p* < .05, ***η***2 = 0.002), a main effect of time point (*F*(1, 2792) = 50.83, *p* < .001, ***η***2 = 0.018) and a main effect of cognitive task (*F*(4.54, 12663.44) = 14.55, *p* < .001, ***η***2 = 0.005). There were also two interactions, one between treatment group and time point (*F*(1, 2792) = 15.30, *p* < .001, ***η***2 = 0.005) (Fig. 4A), and the other between treatment group and cognitive task (*F*(4.88, 13624.05) = 5.20, *p* < .001, ***η***2 = 0.002) (Fig. 4B). The three-way interaction among treatment group, time point, and cognitive task was not significant (*F*(4.88, 13624.05) = 1.64, *p* = 0.15, ***η***2 = 0.00059).

**Figure 4.**
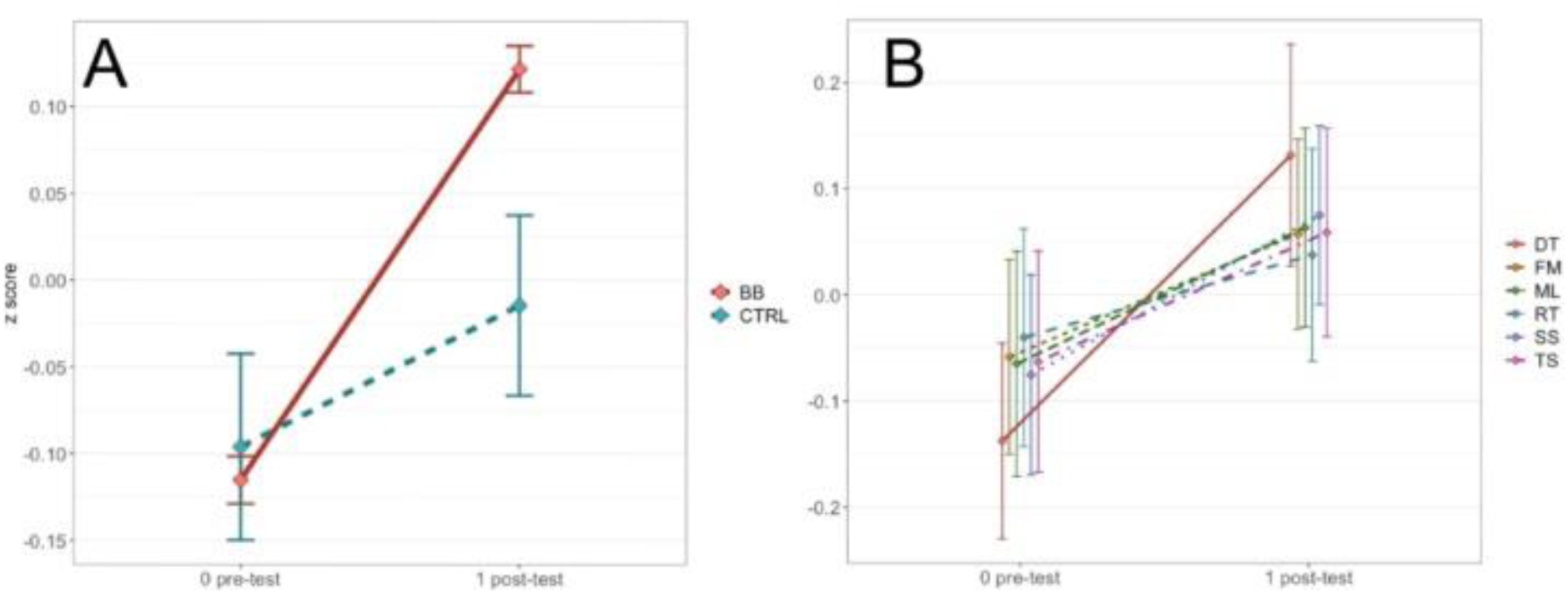
Panel A shows the two-way interaction between time point and treatment group for the in-center program. Panel B shows the two-way interaction between time point and cognitive task for the in-center program.

In the post-hoc analysis, the timeNex point was fixed to either pre-test or post-test. We then tested the two-way interaction between cognitive task and treatment group at each time point. The interaction between cognitive task and treatment group is significant at post-test (*F*(1, 22006) = 25.1, *p* < .001, ***η***2 = 0.001) but not at pre-test (*F*(1, 21775) = 0.49, *p* = .49, ***η***2 = 0.00002). Next, we tested which cognitive tasks had a significant treatment effect between the BB group and the CTRL group at post-test. Among all six tasks, Double Trouble (*F*(1, 3346) = 13.2, *p* < .001, ***η***2 = 0.004), Feature Match (*F*(1, 4287) = 6.86, *p* < .01, ***η***2 = 0.002) and Rotations (*F*(1, 3231) = 7.37, *p* < .01, ***η***2 = 0.002) demonstrated significant treatment effects (Fig. 5).

**Figure 5.**
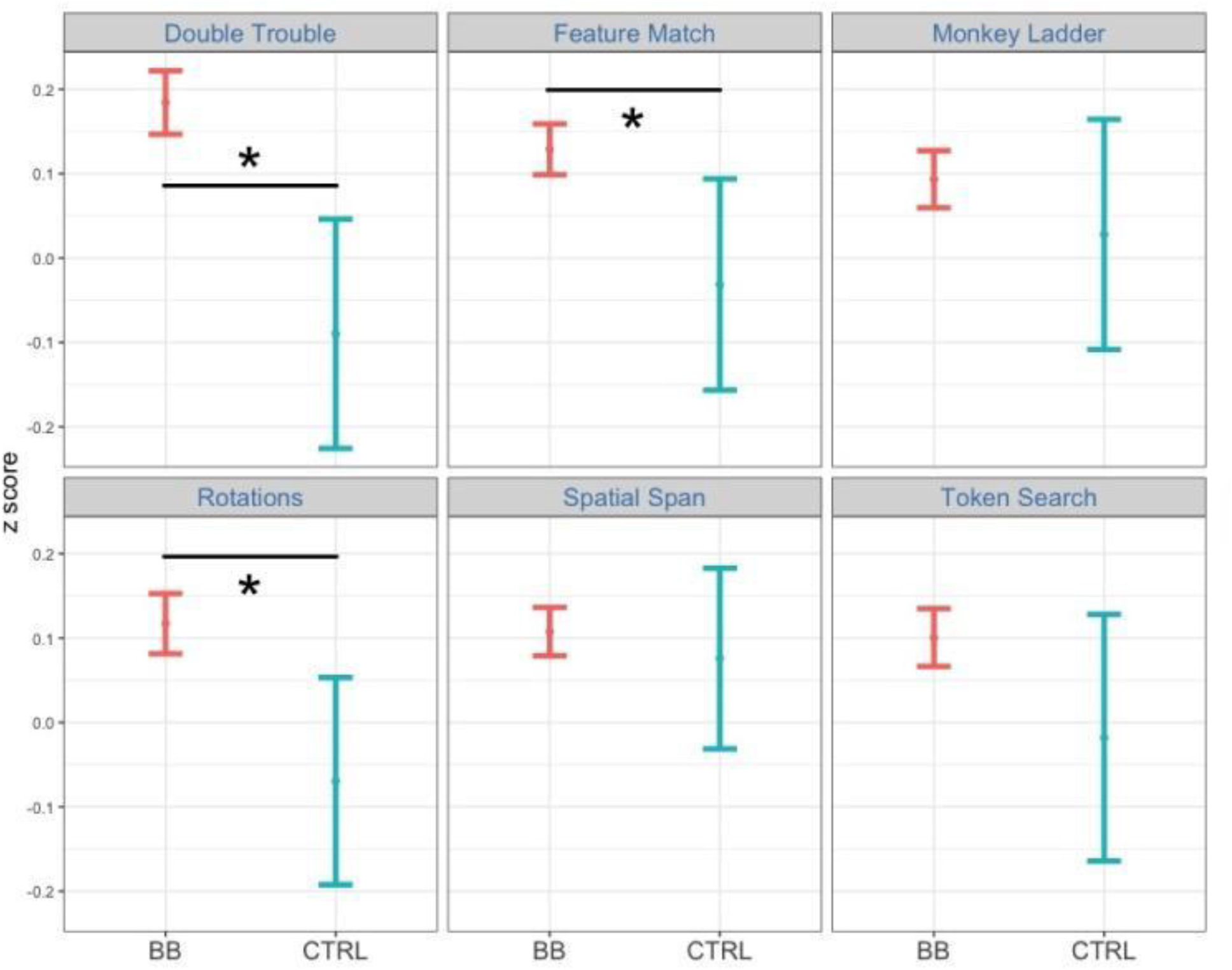
Comparing performance between the BB group and the CTRL group at post-test for the in-center program. Among the six tasks, participants in the BB group outperformed those in the CTRL group in Double Trouble, Feature Match, and Rotations.

In the repeated measures ANOVA assessing test scores from the at-home and the in-center BB group, the homogeneity of variance assumption was met in all but one condition (the Monkey Ladder task at post-test). The main effect of program type was not significant (*F*(1, 2935) = 0.25, *p* = .61, ***η***2 = 0.000086), but the main effect of cognitive task (*F*(4.55, 13363.83) = 43.88, *p* < .001, ***η***2 = 0.015), the main effect of time point (*F*(1, 2935) = 263.46, *p* < .001, ***η***2 = 0.082), and the interaction between cognitive task and time point (*F*(4.88, 14315.43) = 4.77, *p* < .001, ***η***2 = 0.002) were all significant. Furthermore, *p*-values from bootstrapped mean change scores are greater than .025 for all six cognitive tasks (DT, *p* = .19; FM, *p* = .28; ML, *p* = .36; RT, *p* = .043; SS, *p* = .91; TS, *p* = .48), suggesting that for a two-tailed hypothesis, the bootstrapped mean change scores are not significantly different at *α* = 5%. Although it is worth noting that the pre- to post-test change scores in the Rotations task are significantly different between the at-home and the in-center programs at *α* = 10%. This is not surprising considering that in the previous analyses, a significant treatment effect was observed for the in-center program but not for the at-home program. Conversely, the Spatial Span task had a significant treatment effect for the at-home program but not the in-center program. Overall, the at-home and the in-center programs had comparable impacts on cognitive performance, as demonstrated by the BB groups’ test scores.

**Figure 6.**
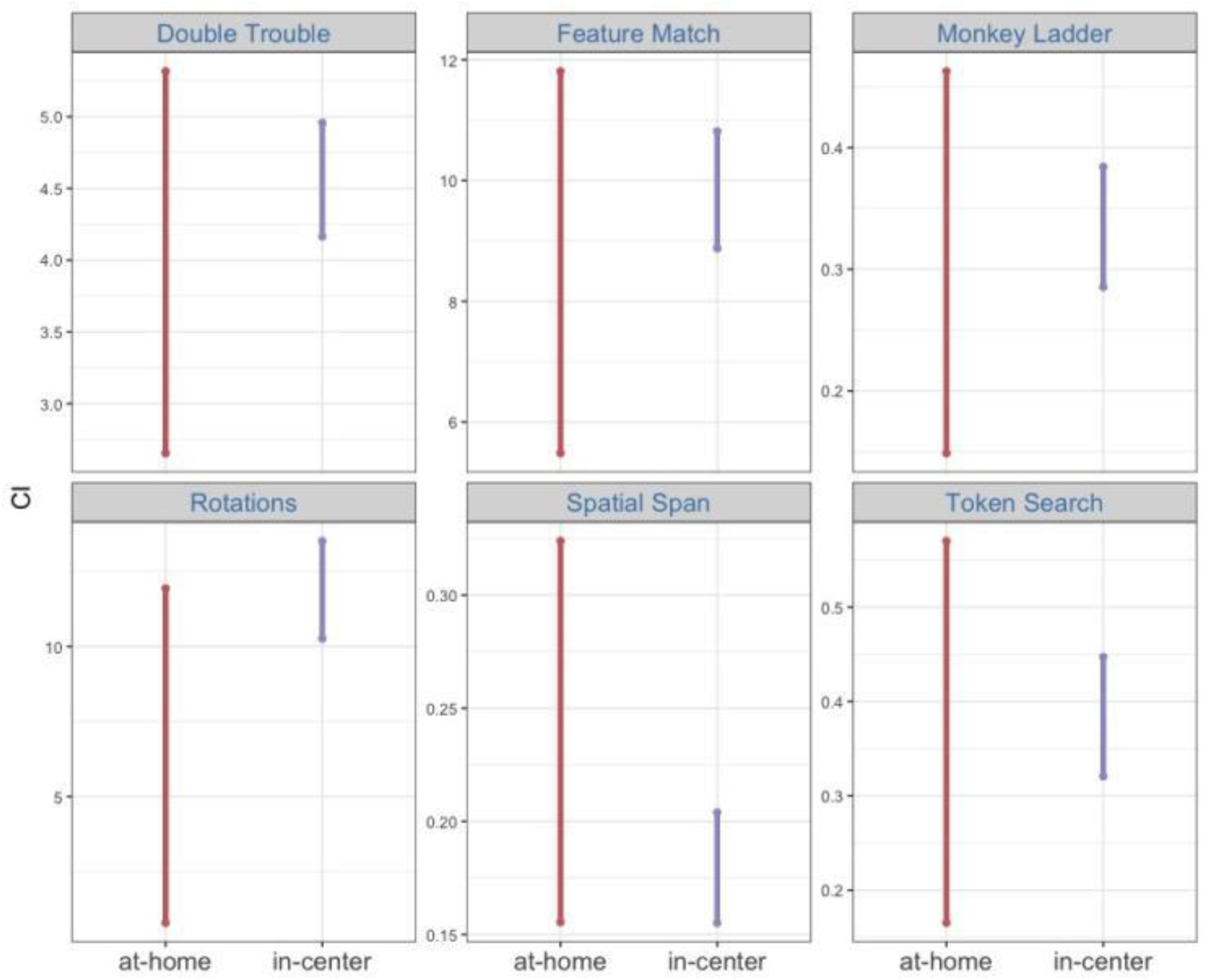
Shown are 95% confidence intervals of bootstrapped means for raw pre-test vs post-test change scores.

## Discussion

In this retrospective evaluation, we reported improved cognitive outcomes of the at-home Brain Balance program — an integrative, multimodal training program — in addressing the needs of students aged 4-17 years with developmental and attentional difficulties. Both at-home and in-center methods of program delivery produced significant treatment effects, with students in the at-home group showing the most notable improvement over controls for measures of attention and response inhibition (Double Trouble), attention and concentration (Feature Match) and short-term memory (Spatial Span), and students in the in-center group showing the most significant improvement over controls in Double Trouble, Feature Match, and a measure of mental rotation and strategy (Rotations). Further, in two analyses directly comparing the at-home versus in-center programs, no significant test score differences were found between the at-home and in-center groups for any of the six cognitive tasks. Overall, the results demonstrate the effectiveness of the Brain Balance program in improving key aspects of cognition including attention, response inhibition, short-term and working memory, and visual-spatial reasoning and strategy in children and adolescents with preexisting developmental and attentional difficulties — even when program delivery is in a virtual at-home format.

A previously published study using a smaller sample size of Brain Balance participants had reported significant improvements from pre-to post-program cognitive performance after participation in the in-center program and significant treatment effects over the control group on specific cognitive tasks (Jackson & Wild, 2021). While the outcomes of Brain Balance participation had previously been investigated in a center-based setting (Jackson & Jordan, 2022, 2023; Jackson & Robertson, 2020; Jackson & Wild, 2021) and in a school setting (Jackson & Glanz, 2023), this is the first study to report Brain Balance participants’ cognitive outcomes in a home-based setting. Using a similar sample size for the at-home program and a much larger sample size for the in-center program, this study replicated and extended the previously published observations of cognitive improvements tested after in-center Brain Balance training and suggested that the at-home Brain Balance program produces cognitive improvements that are comparable to those observed after the in-person center-based Brain Balance program. Collectively, these studies demonstrate the generalizability of the program’s outcomes across different types of settings. Future studies will need to elucidate which aspects of the at-home versus in-center programs may result in greater improvement in some cognitive tasks more than others.

Although virtual delivery of non-medical developmental interventions has not yet been well studied, there is some evidence showing that, for some interventions in children with developmental conditions, both at-home and in-person delivery modalities result in symptom reduction, with no differences between groups that receive at-home versus in-person services (Himle et al., 2012). The benefits of virtual intervention services have been shown to carry over to school, with improvements in student engagement and academic outcomes (Langbecker et al., 2019), and has been recommended to be adopted into future care delivery models for children because of high satisfaction ratings and the convenience and comfort of children receiving services in a familiar home environment (Fairweather et al., 2016; Tenforde et al., 2020). The results presented here on the at-home Brain Balance program are in line with previous studies showing the feasibility and efficacy of pediatric telehealth (Beani et al., 2020; Burke et al., 2015; Cady et al., 2008; Chen et al., 2013; Gloff et al., 2015; K. Myers et al., 2015; K. M. Myers et al., 2007; Sharma et al., 2020) and add to the emerging literature on the value of virtual behavioral therapies or interventions for children with developmental issues (Himle et al., 2012; Singer et al., 2018; Tenforde et al., 2020).

One of the well-documented benefits of telehealth is that it enables provision of care to patient populations residing in underserved areas with limited availability of high-quality, in-person health services (Burke et al., 2015; Gloff et al., 2015; K. M. Myers et al., 2007; Rabatin et al., 2020). Non-medical remote training programs such as the at-home Brain Balance program could similarly serve as a promising way to allow access to childhood development programs in areas with a shortage of in-person centers and services. A recent report from the U.S. Centers for Disease Control and Prevention found a higher prevalence of developmental conditions in children aged 3-17 years living in rural compared to urban areas (Zablotsky & Black, 2020), with these differences being most pronounced for attention-deficit/hyperactivity disorder (ADHD). At the same time, children in rural areas were much less likely to receive intervention or therapy services. Further, pediatric populations with developmental difficulties were shown to be disproportionately affected by closures of in-person educational programs and services during the COVID-19 pandemic (Aishworiya & Kang, 2020; Bentenuto et al., 2021; Lee, 2020; Meireles & de Meireles, 2020). As a training program that is entirely remote and online, the at-home Brain Balance program has the potential to reach large, geographically dispersed populations and may serve as a model for the remote provision of integrative multimodal activities to students with developmental and attentional difficulties who might otherwise go without much-needed support.

## Limitations

The at-home nature of the program requires parental involvement to a greater extent than does the in-center program. Because of potential variability in education levels of our participants’ parents, any instructions to parents are given in plain language that would be simple to understand. However, to what degree families are following these instructions in the intended manner was not tracked in the present study, which may have led to potential differences among participants in implementation of the program. Since there were no significant differences found between participants’ outcomes in the at-home versus in-center programs, the participants were likely adhering to all of the program’s components as intended. However, to fully ensure program fidelity, future studies on the at-home program would need to track and document participants’ adherence to the program.

## Conclusions

In this retrospective evaluation of cognitive test results, we presented novel findings showing improved cognitive performance in children and adolescents after three months of participation in the virtual at-home Brain Balance program. The program was delivered through a telehealth model involving a professional Brain Balance coach and the program’s proprietary app as a digital therapeutic tool within a comprehensive program, in addition to physical exercises and the use of specialized sensory gear, supported by healthy nutrition recommendations. The similar cognitive outcomes found between the at-home and in-center Brain Balance programs suggested that the cognitive effects of virtual training are consistent with those observed for in-person training. Overall, the results demonstrate the effectiveness of the at-home Brain Balance program in improving cognitive functioning in pediatric populations with preexisting developmental and attentional difficulties, including improvements in attention, inhibitory control and memory. This evidence suggests the value of the virtual delivery model when in-person programs are not available or accessible, as well as to supplement routine in-person educational programs and services as needed.

## Data Availability

All data produced in the present study are available upon reasonable request to the authors.

## Acknowledgements

The authors thank Azra Jaferi PhD for editorial direction in scientific writing and publishing, and Joaquin A. Anguera PhD for overall scientific guidance and feedback.

## Author Contributions

This work was carried out in collaboration between both authors. Author RJ designed the study, provided the Brain Balance protocol, and contributed to the manuscript. YM performed the statistical analyses and interpretation of the data, creation of figures and tables, and manuscript writing. Both authors read and approved the final manuscript.

